# Barriers and incentives in the uptake of family planning services: Evidence from the Universal Access to Quality Contraception Project

**DOI:** 10.64898/2026.09.20.26363531

**Authors:** Kanksha Barman, Indrani Gupta, Kali Prosad Roy, Ravi Subbiah

**Affiliations:** Health Policy Research Unit, Institute of Economic Growth, University Enclave, Delhi 110007 India; Charité Center for Global Health, Charité – Universitätsmedizin Berlin, 10117 Germany; Population Services International India Private Limited, 8, Guru Ravidas Marg, Balaji Estate, Kalkaji, New Delhi, Delhi 110019 India

**Author notes:** Corresponding Author: Kanksha Barman.

## Abstract

Population Services International (PSI) set out to demonstrate the engagement of the private sector in increasing access to modern contraceptives through innovative financing strategies, with a focus on marginalised populations, by implementing the SARAL or Universal Access to Quality Contraception (UAQ) project in the city of Agra, Uttar Pradesh. The objective of the UAQ program was to provide a full basket of choices of family planning methods to newly-married women, incentivising them to delay pregnancy for a specified period. A cross-sectional baseline survey was conducted in Agra during the month of June 2022, covering 1240 currently married women. Using baseline survey data, this paper examined the determinants of women’s preference for an additional child, and whether supply-side family planning interventions alone are sufficient to drive adoption of family planning methods. Regression analysis was done to find the determinants of wanting more children and the determinants of the willingness to join the program. The results show that the two most significant variables in determining a woman’s preference for another child are the total number of children she already has, and not having any male children resulting in a strong son preference. The most significant variable in determining a woman’s preference for enrolling herself in the SARAL program is whether she has faced financial constraints in accessing family planning methods, with other determinants being inconvenience of using contraceptive methods, exposure to the government’s outreach about family planning, and preference for a male child. The findings indicate that women’s preferences for an additional child are mostly based on their own desires, and are generally determined by the number and gender of their existing children. However, when it comes to the use of family planning methods, the husbands’ views are a critical factor in making family planning decisions – this would imply that family planning programs need to reach out not just to women, but to their husbands as well. Raising awareness of, and increasing exposure to family planning services is much needed, and programs should incorporate a strong Information, Education and Communication (IEC) component.

## 1. Introduction

One of the major objectives of India’s National Health Policy (2017) is to meet more than 90 percent of the country’s family planning (FP) needs at the national and sub-national level by 2025 [1]. The Indian government’s vision is to provide access to high-quality, comprehensive FP services to all people of reproductive age, with particular emphasis on marginalized groups, by ensuring equitable, affordable, and appropriate contraceptive choices through improved health systems and community engagement within the country’s Universal Health Coverage (UHC) framework by 2030 [2]. Family planning achievements are also central to several of the 2030 Sustainable Development Goals (SDGs), given their close links to gender equity, employment, poverty reduction, and health outcomes [3].

Despite this policy commitment, access to modern contraception remains a pressing challenge, particularly for India’s urban poor. An estimated 24 million people living in urban poverty lack access to FP products [4]; approximately 7.3 million women in urban slums require modern contraceptive methods but are unable to access them, while more than 2 million existing FP users need health system support to continue their current method [5].

At the national level, data from the National Family Health Survey (NFHS) show progress: India’s total fertility rate (TFR) declined from 2.2 in 2015–16 to 2.0 in 2019–20, while the contraceptive prevalence rate (CPR) rose from 54% to 67% over the same period, with modern contraceptive use reaching 56.4% by 2019–20 [6]. However, these national averages mask substantial variation across states.

Uttar Pradesh (UP), home to nearly 17% of India’s population, is a critical state for achieving the country’s FP 2030 vision given its size. According to NFHS-5, UP’s modern CPR stands at just 45%, and its TFR (2.4) remains above the national average. While unmet need for contraceptives in the state declined by 30% between NFHS-4 and NFHS-5, this progress has been uneven across districts – Agra, for example, recorded a decline of only 10% [7].

Access is also uneven: despite substantial investment by donors and institutions in UP’s FP sector, free-of-cost provision remains largely confined to public facilities. Only about 37% of urban women and 60% of rural women access modern contraceptives through the public sector, with the remainder relying on largely unregulated private facilities, medical stores, and shops – leaving many either dependent on the public system or without access to modern methods due to cost and availability constraints.

Further gaps are observed from behavioral indicators: in UP, 16% of women aged 20-24 were married before the legal minimum age of 18, and 3% of women aged 15-19 have already begun childbearing. Birth spacing remains low, with 13% of births occurring within 18 months of a previous birth and 30% within 24 months.

That FP practices remain limited among young married women, despite decades of programmatic investment, raises a central question: what motivates women to adopt family planning, and how does this relate to their preferences for having children or additional children?

Literature directly addressing these questions in the Indian context remains limited, though related studies offer relevant insights. A 2023 study using NFHS-3 and NFHS-5 data identifies a wide set of determinants of preference for additional children, including couples’ age and educational attainment, religion, wealth quintile, residence, age at marriage, son preference, child mortality, age at first intercourse, prior abortion, age at first birth, marriage-to-birth interval, postpartum fecundability, number of daughters, FP knowledge, cultural factors, and media exposure [8].

Evidence from other developing country contexts points to similar patterns. A 2020 study using Demographic and Health Survey data from 32 sub-Saharan African countries found that younger women, women with no formal education (or partners with no formal education), women with no or only one living child, those perceiving six or more children as ideal, women not in paid work, women with limited media exposure or decision-making autonomy, and women in rural or poorer areas were more likely to want more children [9]. Separately, a study of women’s empowerment and fertility preferences in high-fertility Sub-Saharan African countries found that empowerment spanning education, economic independence, and decision-making authority, was significantly associated with a desire for fewer children [10].

Within India, a study of fertility preferences among women in West Bengal found that education, age at marriage, and contraceptive use were significant determinants, with stronger effects in rural areas than urban ones [11]. Son preference, in particular, has been consistently identified as a key factor shaping fertility decisions in India and other developing countries [12].

To address these gaps, Population Services International (PSI) implemented the SARAL, or Universal Access to Quality Contraception (UAQ), project in Agra, Uttar Pradesh, aimed at demonstrating how private-sector engagement – through innovative financing strategies – can expand access to modern contraceptives for marginalized populations. The objective of the UAQ program was to provide a full basket of choices of family planning methods to newly married women, incentivizing them to delay pregnancy for a specified period. For this, a baseline survey was conducted in Agra, UP, by the project partner Population Council during June 2022, collecting data from 1240 currently married non-pregnant women aged 18-49 who had not undergone female sterilization or a hysterectomy.

The UAQ project sought to broaden the range of FP providers and methods available through the private sector, reduce access barriers, address high unmet need, and meet client demand for sterilization by offering a full basket of family planning choices to newly married women, with the goal of incentivizing delayed pregnancy for a specified period.

Using this baseline data, this paper attempts to answer two questions: (a) What determines the preference for a child, or an additional child, among young women? (b) Are supply-side FP interventions sufficient to drive adoption, or do other barriers prevent women from adopting FP practices?

Section 2 presents summary statistics from the baseline survey, covering sample characteristics and knowledge, beliefs, and attitudes toward family planning. Section 3 presents regression results on the determinants of wanting more children and of willingness to join the UAQ program, Section 4 concludes with a summary of findings and key policy recommendations.

## 2. Data, sample and methods

Data for the cross-sectional baseline survey was collected during 1-29 June 2022 in Agra, UP, from a sample of 1240 currently married non-pregnant women aged 18-49 who have not undergone female sterilization or a hysterectomy. Participants were selected through a multi-stage random sampling procedure: urban primary health centres (UPHCs) were randomly selected in the study area, followed by random selection of ASHA (Accredited Social Health Activist) catchment areas within each UPHC, from which eligible women were randomly selected for interview using each ASHA’s eligible-couple register. Data was collected through face-to-face interviews using a structured questionnaire, prepared in English and translated into Hindi, and administered by trained investigators using Computer-Assisted Personal Interviewing (CAPI) on tablets. To minimize bias and encourage honest reporting, interviews were conducted privately, confidentiality of responses was assured, and participants could decline any question. This study was approved by the Sigma Institutional Review Board on 18.05.2022 and the IRB number for the approval is 10096/IRB/21-22. Written informed consent was obtained via signature prior to involvement in the study; where a participant was unable or unwilling to sign, the investigator marked the form to indicate oral consent.

### a. Socioeconomic background

Key social and demographic characteristics of the respondents – age range, caste and religion – are presented in Table 1.

**Table 1:**
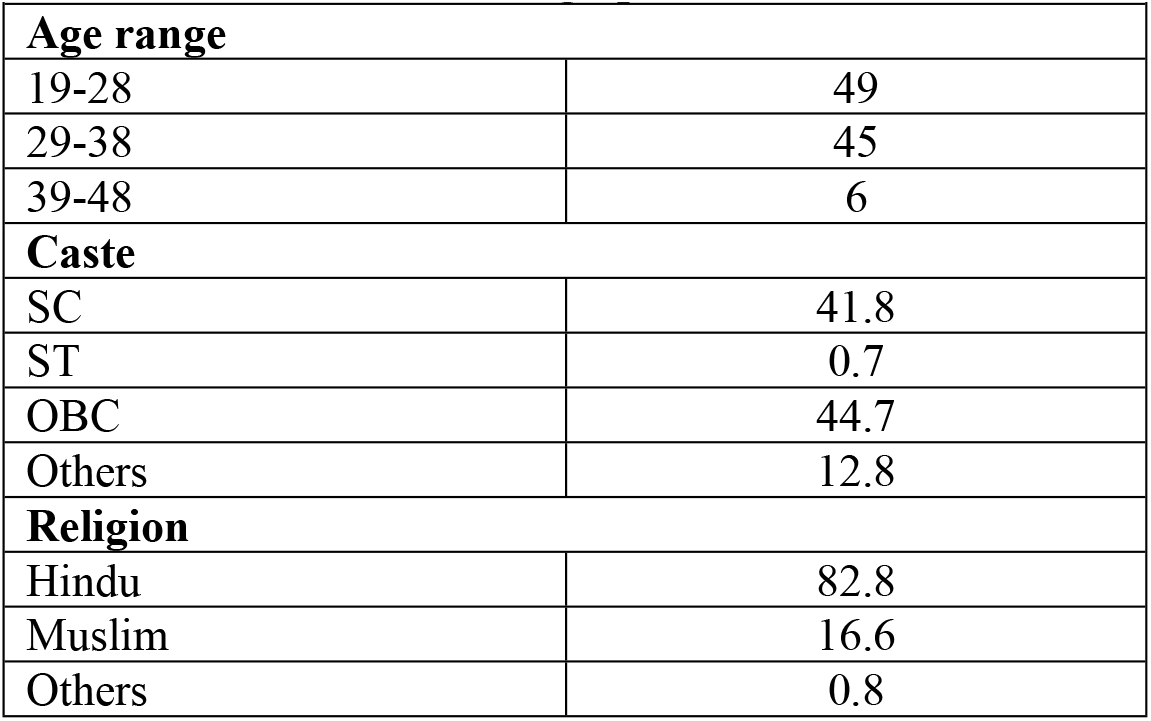
Social and demographic characteristics (%)

The mean age in the sample was 29, with women mostly belonging to the 19-28 or 29-38 age category. As for caste, majority of women belonged to the Scheduled Caste (SC) category (42%) and Other Backward Classes or OBC (45%). Mostly the women were from Hindu religion (83%), with Muslims comprising about 17%.

The mean age at first marriage of the respondents was 19. This is lower than the mean age of marriage among women in India (22.7 years) and in UP (22.5 years), as per data from a 2022 MoSPI report [13]. A majority of women in the sample (73.5%) reported being married in the age range 18-24. Another 21.3% women were married in the age range 10-17 and 5.1% were married over the age of 25 years.

Almost the entire sample of women (99%) had already given birth to a child, with most women having two (42%) or three (27%) children. However, 20% of the women did not have a male child. Only about 24% women reported wanting more children in the future, which we use as a dependent variable in the regression analysis.

As for the educational characteristics of the respondents and their husbands (Table 2), slightly less than a quarter of the sample of women did not have any formal education. This statistic was better for the husbands. Overall, the husbands were better educated than the wives.

**Table 2:**
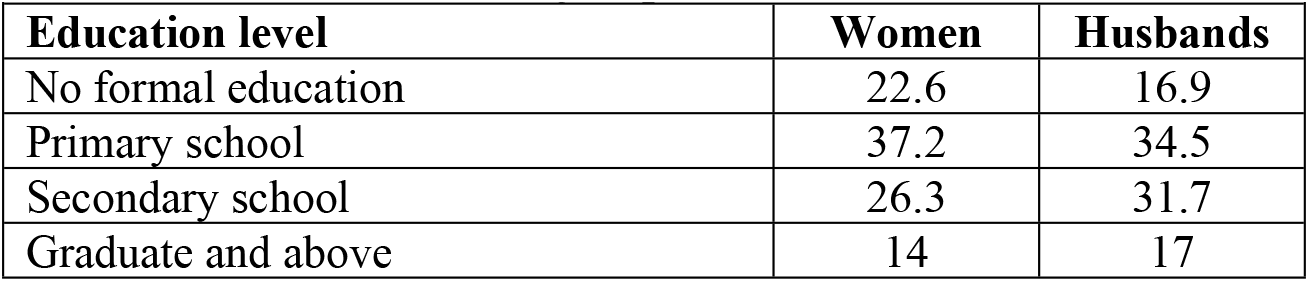
Educational level of respondents and their husbands (%)

| ***Table 2: Educational level of respondents and their husbands (%)*** |  |  |
| --- | --- | --- |
| **Education level** | **Women** | **Husbands** |
| No formal education | 22.6 | 16.9 |
| Primary school | 37.2 | 34.5 |
| Secondary school | 26.3 | 31.7 |
| Graduate and above | 14 | 17 |

Table 3 gives the main or primary occupations of both the respondents, as well as their husbands. The majority of women (81.4%) were homemakers or not engaged in paid work, while 9.1% were engaged in daily wage labour. Among the husbands, daily wage labour was the primary occupation for nearly half (45.1%), while a significant number were also engaged in private jobs (24.2%) or had their own business (26.8%).

**Table 3:** Main occupation of respondents and their husbands (%)

| ***Table 3: Main occupation of respondents and their husbands (%)*** |  |  |
| --- | --- | --- |
| **Occupation** | **Women** | **Husbands** |
| Farmer |  | 0.1 |
| Wage labour | 9.1 | 45.1 |
| Government job | 0.2 | 2.1 |
| Private job | 1.2 | 24.2 |
| Own business | 6.7 | 26.8 |
| Student | 0.3 | 0.3 |
| Homemaker/not working | 81.4 | 1.1 |
| Other | 1.2 | 0.4 |

Table 4 presents the ownership of assets as reported by the respondents. Nearly all (over 98%) the women in the sample reported having an electricity connection, ceiling fan and LPG stove. The majority of women (60-80%) also reported having two-wheelers, color televisions and refrigerators in their homes. Washing machines were owned by over a quarter of women, while personal computers or laptops, four-wheeled vehicles and air conditioners were owned by just 4-6% of respondents. Clearly, the sample comprised individuals who were not very poor, but belonged to lower economic category.

**Table 4:** Ownership of assets (%)

| **Table 4: Ownership of assets (%)** |  |  |
| --- | --- | --- |
| **Asset** | **Yes** | **No** |
| Electricity connection | 98.9 | 1.1 |
| Ceiling fan | 98.2 | 1.8 |
| LPG stove | 98.2 | 1.8 |
| Two-wheeler | 63.2 | 36.8 |
| Color TV | 79.2 | 20.8 |
| Refrigerator | 75.3 | 24.7 |
| Washing machine | 28.9 | 71.1 |
| Personal computer/laptop | 6 | 94 |
| Car/jeep/van | 6.1 | 93.9 |
| Air conditioner | 4.6 | 95.4 |

### b. Family planning: service use, methods, beliefs and attitudes

Table 5 presents responses on the current use of FP methods, method mix, and awareness of side effects. A very high percentage – almost 78% – of the respondents reported that they are currently using FP methods, of which 70% reported using modern methods while 30% reported using traditional methods. More than 60% of the respondents reported being aware of side effects when they first started using the FP method.

**Table 5:** Use of family planning methods (%)

| **Table 5: Use of family planning methods (%)** |  |
| --- | --- |
| **Currently using family planning method** |  |
| Yes | 77.7 |
| No | 22.4 |
| **Type of family planning methods being used** |  |
| Modern methods | 69.9 |
| Traditional methods | 30.1 |
| **Awareness of side effects when FP method was first used** |  |
| Yes | 60.2 |
| No | 39.8 |

Table 6 presents women’s awareness about FP methods, which in turn determines current or past use of FP methods.

**Table 6:** Awareness of FP methods (%)

| *Table 6: Awareness of FP methods (%)* |  |  |
| --- | --- | --- |
| **FP methods** | **Yes** | **No** |
| Female sterilization | 96.5 | 3.5 |
| Male sterilization | 75.4 | 24.6 |
| IUCD/PPIUCD | 94.7 | 5.3 |
| Injectables | 92.1 | 7.9 |
| Pill | 94.3 | 5.7 |
| Condom/Nirodh | 98.2 | 1.8 |
| Emergency contraception | 8.6 | 91.4 |
| Lactational amenorrhea method | 14.2 | 85.8 |
| Rhythm method | 85.8 | 14.2 |
| Withdrawal | 81.7 | 18.3 |
| Other methods | 7.2 | 92.8 |

Awareness of FP methods – especially modern methods – was almost universal in the sample. Almost all women knew about condom as a FP method (98%). However, for other modern methods as well, the awareness was over 90%. Awareness of female sterilization (96.5%) was much higher than awareness of male sterilization (75.4%), which is not surprising given the push for female sterilization in Indian FP programs. Among traditional methods, the rhythm method (85.8%) and withdrawal (81.7%) were known by most women, while awareness of the lactational amenorrhea method (LAM) was low (14.2%). Emergency contraceptive pills also had very low awareness (8.6%).

Table 7 presents responses to selected questions relating to knowledge, attitudes, and beliefs about overall understanding or attitudes about family planning, including the source from where the women could avail services. Knowledge of availability of FP methods is generally high with 93.3% of women reporting that they knew places in their area where FP methods are available. A majority of women had a positive attitude towards FP with 77% reporting that use of FP methods improves women’s health and only 31.5% reported feeling that using FP methods was a hassle.

**Table 7:** Knowledge, attitudes and beliefs about FP methods (%)

| *Table 7: Knowledge, attitudes and beliefs about FP methods (%)* |  |  |
| --- | --- | --- |
| **Knowledge** | **Yes** | **No** |
| You know places in your area where FP methods are available | 93.3 | 6.7 |
| FP methods are easily available in your area | 90.7 | 9.3 |
| FP methods are always available in your area when you need them | 88.1 | 11.9 |
| **Attitude** | **Yes** | **No** |
| Using FP methods improves women's health | 77 | 23 |
| Using FP methods is a hassle for me | 31.5 | 68.5 |
| **Belief** | **Yes** | **No** |
| Use of FP methods spoils the uterus | 30.4 | 69.6 |
| Use of FP methods makes a woman physically weak | 35.3 | 64.7 |
| **Social support** | **Yes** | **No** |
| Choice to use FP methods is up to your husband | 62.7 | 37.3 |
| You believe that your in-laws don't like you to use FP methods | 21.2 | 78.8 |
| Your husband doesn't like to talk about FP | 25.4 | 74.6 |
| If you had any problems with family planning method, your husband would help you | 97.9 | 2.1 |

Overall, negative impression of FP was present in about slightly more than one-third of the sample women, which is not a very low and can act as a deterrent to adopting FP methods.

Social support from husbands in using FP methods was almost universal with almost 98% of women reporting that their husbands would help them if they had a problem while using any FP method. Also, support from in-laws was quite high as well, with only 21.2% reporting that their in-laws did not want them to use FP methods. However, and most importantly, a majority of women (62.7%) also said that the choice to use FP methods was up to their husbands. About a quarter of women also reported that their husbands did not like to discuss FP issues with them.

A key factor for wanting to enroll in FP programs such as the SARAL program is whether women face financial constraints in accessing healthcare and specifically, FP services. Table 8 presents the percentage of women who reported financial constraints in the last 12 months.

**Table 8:** Financial constraints during the last 12 months (%)

| ***Table 8: Financial constraints during the last 12 months (%)*** |  |  |  |  |
| --- | --- | --- | --- | --- |
| **Response** | **Purchase of medicines** | **Visiting private doctor** | **Purchase of FP product** | **Visiting private doctor for FP services** |
| Yes | 36.8 | 34.2 | 12.7 | 9.5 |
| No | 59 | 61.1 | 69.9 | 72.7 |
| Not needed | 4.2 | 4.7 | 17.4 | 17.8 |

While more than one-third (34-37%) reported financial constraints in the purchase of medicines or visiting a private doctor, fewer women (about 13%) reported financial constraints in the purchase of FP products and less than 10% of women reported financial difficulties in consulting a private doctor for FP services.

Consumption of media and advertising has often been mentioned as an important factor in raising awareness about interventions. Very few women (5-7%) reported reading newspapers or magazines whether daily or once in a while, and less than 3% reported listening to the radio. However, almost 61% of women reported watching television every day and only about 20% of women did not watch television at all. About 40% reported owning a smartphone, while others reported that they could access the smart phones of some other family member. This is quite high, and holds the potential for access to information. However, given the education levels and consumption of information from media, it is not clear whether mere ownership could lead to transfer of important information about interventions like FP programs.

A very important variable is access to government healthcare workers who can give information and advice about FP methods. About 39% women reported that they had been approached by or had come into contact with healthcare workers in the past 3 months, with whom they were able to discuss FP concerns, while 61% of women reported that they had not. Of those women who reported yes, 89% said they discussed FP concerns with an ASHA, 40% with a doctor/nurse from a private hospital/clinic, 20% with an ANM, 15% with a doctor/nurse from a government hospital/clinic and 12% with an *anganwadi* worker.

The respondents were also asked whether they had seen any FP-related advertisements in the past 3 months. About 30-33% of the respondents had seen posters or wall paintings advertising FP, while 14.5% had seen billboards, and about 5% reported seeing leaflets on FP.

A key variable in the analysis is the willingness to enroll in the SARAL program. About 70% of the women agreed to join the program, while 30% refused to join. Tables 9 and 10 present the reasons for their choice to enroll in the program. It may be noted that multiple responses were received from the respondents.

**Table 9:** Reasons for wanting to join the SARAL program (%)

| ***Table 9: Reasons for wanting to join the SARAL program (%)*** |  |
| --- | --- |
| Access to FP services free of cost | 45.1 |
| Access to FP services nearby | 15.6 |
| Need FP methods | 11.8 |
| Access to FP services in private sector | 9.7 |
| Ease of access | 6.3 |
| Guaranteed access to services | 5.7 |
| Better quality of services | 1.6 |
| Access to information about FP methods and side effects | 2.1 |
| Don't know | 2.1 |

**Table 10:** Reasons for not wanting to join the SARAL program (%)

| ***Table 10: Reasons for not wanting to join the SARAL program (%)*** |  |
| --- | --- |
| FP services not needed at this time | 46.9 |
| Already using long-term FP method or husband brings condoms when needed | 7.4 |
| Planning to have a child soon | 7.4 |
| FP methods already available free of cost | 4.9 |
| Don't like modern FP methods | 4.9 |
| Need to ask husband/in-laws | 4.9 |
| Worry of side effects or method failure | 4.9 |
| Plan to get sterilized | 3.7 |
| Infertile | 2.3 |
| Husband is away or less frequent sex | 1.7 |
| Don't have faith in such a program | 1.7 |
| Husband/in-laws have said no | 1.4 |
| Don't have financial constraints | 0.9 |
| Prefer private hospital | 0.3 |
| Don't know | 6.9 |

The most common reason for wanting to enroll in the program was access to FP services free of cost, as reported by almost half (45.1%) of the women. Other important reasons were: need for FP methods and services (11.8%), access to FP services nearby through the program (15.6%), access to FP services in the private sector (9.7%) – again through the program, and ease of access or saving time (6.3%). Other reported reasons were guaranteed access to services, better quality of FP services, and access to information about FP methods and their side effects.

The most common reason (47%) for declining to enroll in the SARAL program (Table 10) was a lack of felt need for FP services at that time. Several other reasons were mentioned like planning for a child, dislike for modern methods, requiring permissions from husbands/in-laws etc.

## 3. Results: Regression analysis

### a. Determinants of preference for additional child

A probit regression was used to analyze the determinants of additional child preference among the respondents.

Education [14] and economic empowerment [15] are often cited as key variables that determine a woman’s choice. We use education, work status, and economic status of household as three key variables that determine additional child preference. Age is used to account for possible decline in preference among older women. Age squared is used to account for non-linearity. Husbands often are the main decision-makers [16], and therefore, we use the husband’s education as another important variable. We also use age at first marriage, and whether she has any male child/children as additional variables. The last is important in the context of male preference among families [12].

Age, age^2^, woman’s education, husband’s education and age at first marriage are used as continuous variables. Awareness depicts women’s knowledge of modern methods of family planning as given in Table 6, and takes a value of 1 if the response was ‘yes’ to being aware of condoms, contraceptive pills, IUCDs, injectables and female sterilization. Hindu, SC/ST and OBC are taken as presented in Table 1. The variable on economic status is built from Table 4 on ownership of assets and conceptualized as poor and non-poor, with those who reported having only an electricity connection, ceiling fan, LPG stove, and two-wheeler being categorized as poor. Finally, no male children and the total number of children are constructed from data on the number of living children, both male and female, as reported by the respondents.

Table 11 presents the probit results with the change in probabilities (instead of the probit coefficients) and their level of significance.

**Table 11:** Probit analysis of the determinants of wanting more children. Dependent variable: Whether want more children (1=want, 0 = don’t want)

| **Table 11: Probit analysis of the determinants of wanting more children** |  |
| --- | --- |
| *Dependent variable: Whether want more children (1=want, 0 = don't want)* |  |
| **Variable** | **Coefficient** |
| Age | -0.053* |
| Age<sup>2</sup> | 0.000~0 |
| Awareness | 0.024 |
| Hindu | -0.087* |
| SC/ST | 0.085* |
| OBC | 0.086* |
| Women's education | 0.004 |
| Husband's education | 0.002 |
| Economic status | -0.017 |
| Age at first marriage | 0.018** |
| Have no male child | 0.614*** |
| Total children | -0.225*** |
| N = 1240, Pseudo R<sup>2</sup> = 0.49<br>LR chi<sup>2</sup>(14) = 731.57<br>Prob > chi<sup>2</sup> = 0.0000<br>Log likelihood = -387.4 |  |
| Note: * significant at 5%, ** significant at 1% and *** significant at 0.1% |  |

The results indicate that two variables that are most significant in determining a woman’s preference for another child are the total number of children she already has, and not having any male children - the latter showing a strong male preference for children. Women with no male children have a 61% higher probability of wanting a child. On the other hand, an additional child reduces the probability by about 23%. Age at first marriage is important as well, and at lower levels of significance so are the variables age, social status, and belonging to a Hindu family.

However, ‘wanting’ another child does not necessarily mean being able to use FP methods in keeping with her additional child preference. Table 12 presents the common social barriers that might hinder a woman’s ability to use FP methods.

**Table 12:** Family barriers in use of FP methods (%)

| **Table 12: Family barriers in use of FP methods (%)** |  |  |
| --- | --- | --- |
| **Statement** | **Yes** | **No** |
| You can use a family planning method even if your husband does not want you to | 16.4 | 83.6 |
| You can use a family planning method without your husband knowing | 12.2 | 87.8 |
| You can use a family planning method even if your mother-in law does not want you to | 79.7 | 20.3 |
| You can continue to use a modern method of family planning even if people in your community find out | 93.9 | 6.1 |

While a majority of the women did not seem to take into account the preferences of their mothers-in-law or the community at large, they were definitely greatly influenced by the opinions of their husbands as far as use of FP methods were concerned. About 84% stated that they will not be able to use a contraceptive method if their husbands do not want to and 88% stated that they will be unable to use a method without their husband’s knowledge.

### b. Determinants of the willingness to join SARAL program

While the various bi-variate tabulation of summary statistics in the previous sections indicated several possible determinants of why women might want to join the SARAL program, a multivariate analysis is required to understand better the major reasons for the willingness to join such a program.

In addition to age and education, other variables used are financial constraints in accessing healthcare, media habits, whether the women have seen any advertisements relating to family planning, whether they have discussed family planning concerns with government health workers, and their beliefs on family planning.

Financial constraints, family planning beliefs, FP being the husband’s choice, media habits, advertisements about family planning and whether able to discuss FP concerns with government health workers are all variables previously discussed.

Table 13 presents the probit results with the change in probabilities (instead of the probit coefficients) as before, and their level of significance.

**Table 13:** Probit analysis of the determinants of willingness to join the SARAL program. Dependent variable: Whether willing to enroll in the SARAL program (1=will join, 0 = won’t join)

|  |  |
| --- | --- |
| **Variable** | **Coefficient** |
| Age | 0.06** |
| Age<sup>2</sup> | -0.001** |
| Awareness | -0.0* |
| Women's education | 0.004* |
| Husband's education | 0.001 |
| Financial constraints | 0.22*** |
| Find FP a 'hassle' | -0.08** |
| FP is the husband's choice | -0.04* |
| Have no male child | -0.08** |
| Total children | 0.03* |
| Media habits | 0.02 |
| Seen advertisements about FP | 0.005 |
| Talked to govt HW about FP | 0.053** |
| N = 1240, Pseudo R<sup>2</sup> = 0.067<br>LR chi<sup>2</sup>(14) = 99.59<br>Prob > chi<sup>2</sup> = 0.0000<br>Log likelihood = -688.1 |  |
| Note: * significant at 5%, ** significant at 1% and *** significant at 0.1% |  |

The most significant variable in determining a woman’s preference for enrolling herself in the SARAL program is whether or not she has faced financial constraints in accessing FP methods in the last year – a woman with such a constraint has a 22% higher probability of joining the program. The other important results are that the probability of joining the program decreases by around 8% if women think that use of FP is a hassle. Exposure to the government’s outreach about FP increases the probability of joining the program by 5%, male child preference decreases the probability of joining the program by about 8%, and belief that FP is the husband’s choice decreases the probability of joining by 4%.

Other significant variables are age and age^2^, but age^2^ has a negative sign, indicating that older women are more likely to want to enroll, but only up to a certain age, after which the likelihood of enrolment decreases. The regression results also show that media habits, awareness of FP methods, and seeing advertisements relating to FP are not significant, but interacting with government health workers was significant with a positive sign.

## 4. Summary and conclusions

The results indicate that women’s preferences for an additional child are mostly based on their own preference, and are generally determined by how many children they currently have and whether they already have had a male child. However, the variable on the views of husbands is the most important determinant of use of FP methods. The husbands’ opinions play a key role with only about 16% women reporting that they would feel confident in using FP methods if their husbands did not want them to, and almost 84% women stating that they would not be able to use a contraceptive method without their husband’s knowledge.

This would seem to imply that a woman may not be able to translate her preferences about children to her preference about FP, the latter being largely determined by her husband’s preferences and beliefs. This also indicates that standard FP programs that are unable to influence the variables that determine a woman’s agency are likely to succeed only to a limited extent, and only where the woman is not constrained by internal family dynamics, especially her husband’s choices. For a large number of women who do depend on their husband’s preferences, it will be critical to reach out to the husbands via FP programs, to change awareness, attitude, and beliefs.

This finding is consistent with existing evidence from sub-Saharan Africa, where women’s empowerment and financial constraints have similarly been shown to shape fertility preferences and family planning uptake [10]. The strong influence of husbands’ preferences on FP use also echoes broader evidence on gender inequality in household decision-making in developing countries [12]. Similarly, the centrality of male-child preference as a driver of continued childbearing has been documented in other South Asian and sub-Saharan African contexts [9, 11], suggesting this pattern reflects a wider regional dynamic.

Also, the results indicate that financial constraints are the most important reason for joining programs that offer low-cost or zero-cost FP products. However, exposure to existing programs is very critical too, because that raises awareness and familiarity with FP features and services. The results also indicate that government outreach influences a woman’s choices about joining FP programs, and thus increasing outreach programs will remain a key intervention of any FP program. Some women still think that FP is cumbersome – a view that can be alleviated with greater and proper exposure to information and communication.

This study has some limitations. First, given its cross-sectional design, the findings reflect associations rather than causal relationships. Second, data on family planning knowledge, attitudes, and practices were self-reported on sensitive topics, which may be subject to social desirability or recall bias, although efforts were made to minimize this through private interviews and assured confidentiality. Finally, information on the husband’s attitudes and role in decision-making was collected only from the wife, without a direct interview with the husband.

The study findings suggest that, with properly designed and subsidized FP services that can reduce OOPE of women on contraceptives and related services, the uptake of FP services may be increased, but will have to be accompanied by a strong Information, Education and Communication (IEC) component, that reaches both the wives and the husbands. Whether the private sector or the government does that is not central to this view, though the government is best suited to expand the IEC component. Both sectors can be present in offering quality and affordable products, but the long-time presence of the government FP program in India has certainly had a significant impact on the accessibility and availability of interventions. The gaps that exist can be filled by the government either solo or in partnerships with the private sector.

## Data Availability

The anonymized dataset underlying the findings of this study is available in the Harvard Dataverse repository, DOI: https://doi.org/10.7910/DVN/XX6FJA

## Acknowledgements

The authors would like to acknowledge colleagues at the Population Council, New Delhi, and Devmani Upadhyay at the Institute of Economic Growth, Delhi.

## Notes

### Competing Interest Statement

The authors have declared no competing interest.

### Author Declarations

Sigma Institutional Review Board, approval #10096/IRB/21-22

